# The Joint Impact of COVID-19 Vaccination and Non-Pharmaceutical Interventions on Infections, Hospitalizations, and Mortality: An Agent-Based Simulation

**DOI:** 10.1101/2020.12.30.20248888

**Authors:** Mehul D. Patel, Erik Rosenstrom, Julie S. Ivy, Maria E. Mayorga, Pinar Keskinocak, Ross M. Boyce, Kristen Hassmiller Lich, Raymond L. Smith, Karl T. Johnson, Julie L. Swann

## Abstract

**Background:** Vaccination against SARS-CoV-2 has the potential to significantly reduce transmission and morbidity and mortality due to COVID-19. This modeling study simulated the comparative and joint impact of COVID-19 vaccine efficacy and coverage with and without non-pharmaceutical interventions (NPIs) on total infections, hospitalizations, and deaths.

**Methods:** An agent-based simulation model was employed to estimate incident SARS-CoV-2 infections and COVID-19-associated hospitalizations and deaths over 18 months for the State of North Carolina, a population of roughly 10.5 million. Vaccine efficacy of 50% and 90% and vaccine coverage of 25%, 50%, and 75% (at the end of a 6-month distribution period) were evaluated. Six vaccination scenarios were simulated with NPIs (i.e., reduced mobility, school closings, face mask usage) maintained and removed during the period of vaccine distribution.

**Results:** In the worst-case vaccination scenario (50% efficacy and 25% coverage), 2,231,134 new SARS-CoV-2 infections occurred with NPIs removed and 799,949 infections with NPIs maintained. In contrast, in the best-case scenario (90% efficacy and 75% coverage), there were 450,575 new infections with NPIs maintained and 527,409 with NPIs removed. When NPIs were removed, lower efficacy (50%) and higher coverage (75%) reduced infection risk by a greater magnitude than higher efficacy (90%) and lower coverage (25%) compared to the worst-case scenario (absolute risk reduction 13% and 8%, respectively).

**Conclusion:** Simulation results suggest that premature lifting of NPIs while vaccines are distributed may result in substantial increases in infections, hospitalizations, and deaths. Furthermore, as NPIs are removed, higher vaccination coverage with less efficacious vaccines can contribute to a larger reduction in risk of SARS-CoV-2 infection compared to more efficacious vaccines at lower coverage. Our findings highlight the need for well-resourced and coordinated efforts to achieve high vaccine coverage and continued adherence to NPIs before many pre-pandemic activities can be resumed.

## Introduction

SARS-CoV-2 vaccines will play a major role in achieving sufficient population immunity to end the COVID-19 pandemic.^1^ In May 2020, the U.S. Department of Health and Human Services set a goal to administer 300 million doses of COVID-19 vaccine by January 2021.^2^Further, with an estimated 16 billion doses required to meet global demand, mass vaccination campaigns across the world will be unprecedented.^3^ In December 2020, the Pfizer-BioNTech^3^ and Moderna^4^ mRNA vaccines were authorized for emergency use, with more expected to follow in 2021.^4-7^

While the primary endpoint of COVID-19 vaccine trials is efficacy to prevent severe disease and death, particularly among vulnerable populations, these vaccines are expected to reduce transmission and contribute to the population immunity required to end the pandemic.^8^Recent mathematical modeling suggests vaccine distribution and uptake need to be prioritized to maximize the benefit of a highly efficacious vaccine.^8-10^ Bartsch, et al. estimated that the U.S. population would require at least 75% coverage with a vaccine efficacy of 70% to reduce the epidemic peak by >99% without other interventions.^9^

Considering the complexities of large-scale vaccine production, distribution, storage, and uptake, achieving high coverage will be challenging.^9^ Therefore, critical questions remain regarding the need to continue non-pharmaceutical interventions (NPIs), such as physical distancing and face mask usage, as the public is vaccinated over time.^8, 11^ Models of COVID-19 transmission that capture complex and heterogeneous interactions between individuals are needed to understand important dynamics between vaccination strategies and other behavioral interventions across diverse populations.

Using an integrated compartmental disease transmission model and agent-based simulation,^12-14^ we estimated the impact of vaccination across hypothetical yet realistic scenarios of vaccine efficacy and population coverage on SARS-CoV-2 infections and COVID-19-related hospitalizations and deaths. Simulating vaccine distribution in North Carolina, we compared the impact of efficacy and coverage scenarios with NPIs maintained and removed concurrently with vaccine distribution. To investigate differences between subpopulations, we stratified the results by race-ethnicity groups and urban, suburban, and rural communities. These objectives were informed by state and local public health decision makers involved in COVID-19 response.

## Methods

### Study Design

We employed an agent-based stochastic network with a susceptible-exposed-infectious-recovered (SEIR) framework for the transmission and progression of SARS-CoV-2 infections.^12-14^ Agent-based models with an SEIR framework have been widely used and accepted for modeling disease spread while considering population behavior.^15-17^ In the SEIR portion of the model, an individual can be in one of the following states at any given time: susceptible, exposed, pre-symptomatic infectious, asymptomatic infectious, symptomatic infectious, hospitalized, and recovered (Figure 1a). Key model assumptions include (i) only hospitalized individuals can transition to death, and (ii) only symptomatic individuals can become hospitalized. Per Keskinocak, et al.,^13^ we simulated heterogeneous mixing and interactions among agents, where daily transmission occurs within households, workplaces and schools, and communities (census tracts), with day and night differentiation in interactions (Figure 1b). The network of agents was constructed using U.S. Census tract-level data on household size (numberof adults and children) and age groups (0-4, 5-9, 10-19, 20-64, and 65+ years old). Model parameters include the basic reproductive number (R0=2.4),^18-20^ asymptomatic and symptomatic transmission coefficients,^18, 21^ and age-specific hospitalization and mortality rates.^22^ Diabetes was included as a risk factor for hospitalization conditional on an agent’s age and race as derived from statewide prevalence data.^23^ Model details and parameter values are provided in Supplement 1. This study was determined to be exempt from IRB review by the UNC Office of Human Research Ethics.

**Figure 1.**
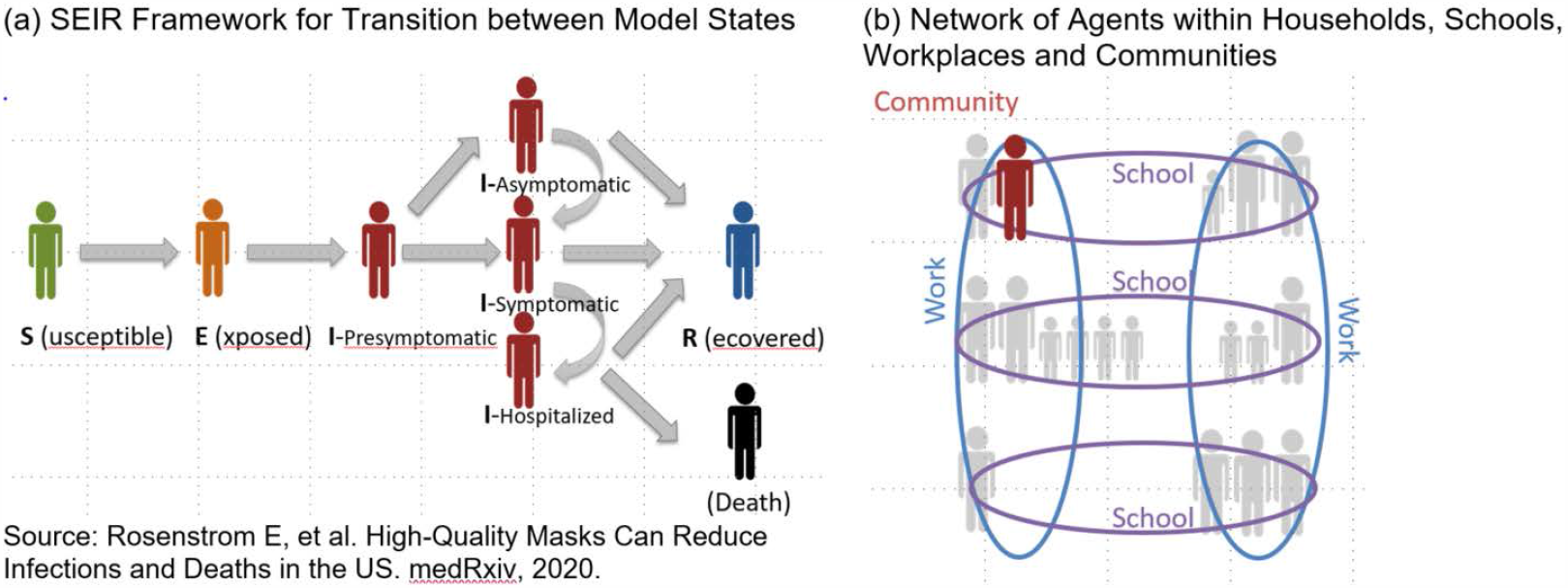
Agent-Based Model Framework and Structure.

### Study Population

The State of North Carolina (NC) was selected as a representative region, with over 10 million people and diversity across age, race/ethnicity, and socioeconomic groups. According to U.S. Census estimates, NC has 18% of persons 65 years or older, 20% Black or African American, 7% Hispanic or Latino, and 19% living in poverty.^24^ Of the 100 NC counties, 80 are rural with an average population density of ≤250 people per square mile.^25^ As of December 21, 2020, NC is ranked 38th in the country by deaths per capita with about 60 total deaths per 100,000 residents.^26^ A stay-at-home order from the Governor’s office was in place from March 24 through May 8, 2020, and face masks have been required in indoor public settings since June 24, 2020. As of August 24, 2020, K-12 schools have been open for both reduced occupancy in-person and remote learning with local school districts determining the operating model, most of which are hybrid.^27^

### Agent-Based Simulation

The NC population of 10,490,000 consisting of 2,195 census tracts was modeled with 1,017,720 agents. The simulation was seeded (“Day 1”) with cumulative infections as of March 24, 2020,^28^ where the initial lab-confirmed cases were multiplied by a factor of 10 to account for limited testing and underreporting^29^ to approximate the true number of infections. This number was then scaled to the number of agents in the simulation. The simulation spanned 18 months (548 days) and was calibrated and validated using the number of lab-confirmed cases and reported hospitalizations and deaths as of November 1, 2020. County-specific cases were randomly distributed to households across the county’s census tracts proportionally to the tract population. For each simulated scenario, 45 replications were performed. A different seed population (i.e., agent demographics and who they interact with and where) was selected for each replication and accounts for the variation across replications. For subgroup analyses, simulation results were stratified by race/ethnicity (Non-Hispanic White, Non-Hispanic Black/African American, and Hispanic). Results were not reported by other race groups like Asian, Native American, and multiracial because of small denominators. We also stratified by urban, suburban, and rural census tracts defined by rural-urban commuting area (RUCA) codes (1-2, 3-5, and 6-10, respectively).

### COVID-19 Vaccination

In the simulations, vaccines were distributed uniformly to adults (20 years and older) over a 6-month period, independent of previous disease or age. We simulated vaccination scenarios in which the distribution started when the population had a cumulative infection prevalence of 10% (Day 213). Three vaccine coverage scenarios reach 25%, 50%, and 75% at the end of the 6-month distribution period. Initial supplies were assumed to be low, followed by increasing amounts with 50% of the final coverage distributed in the first 4 months and 50% distributed in the last two months. Coverage represents the proportion of adults receiving the vaccine, which is a function of both vaccine availability and uptake, although these factors were not modeled individually. In our study, vaccine efficacy was modeled as the probability that a vaccinated susceptible agent directly transitions to the recovered state. We assumed the vaccine confers immunity immediately. Vaccine efficacy scenarios tested were 90%, based on preliminary estimates of vaccine efficacy from Phase 3 trials, and 50 %, based on the minimum efficacy threshold established by the FDA. We simulated the two efficacy levels and three coverage levels, resulting in six vaccination scenarios (A-F); the “best case” being (A) 90% efficacy, 75% coverage, and the “worst case” being (F) 50% efficacy, 25% coverage. In addition, we simulated a “no vaccine” scenario (G) to provide context.

### Non-Pharmaceutical Interventions

Limited mobility (i.e., physical distancing) was estimated with SafeGraph mobility data^30^aggregated by census tract RUCA code and income quartile. Data reflected the stay-at-home order in place from March 24 to May 8, 2020 for non-essential workers as well as reductions in distancing over time. We also modeled voluntary isolation and quarantine, where an agent with symptomatic infection isolated at home with all members of the household until the agent recovered. Based on mobility data and computational experiments, this occurred with 60% probability initially and then decreased by 15 percentage points each week until 15%, where it remained for the duration of the simulation.

Face mask usage among agents began at 0% and increased at a constant rate to 70% when face masks were mandated statewide in June. Masks were assumed to reduce susceptibility to infection and infectivity to others by 50% each. Since schools closed in March and moved to remote instruction, there were no K-12 school interactions until August 24 when schools were allowed to open under a hybrid policy that limited the total number of students to ensure distancing. After August 24, agents were split into two equal groups where one group attended school on even dates and the other attended on odd dates.

We simulated vaccination scenarios (A-F) with these NPIs maintained (A1-F1) and removed (A0-F0). When NPIs were maintained, they remained at pre-vaccination levels throughout the simulation. When NPIs were removed, workplace and community interactions including schooling returned to normal and agents no longer isolated or quarantined immediately at the midpoint of the 6-month vaccine distribution period. Face mask usage decreased at a rate proportional to the number of agents vaccinated until the end of vaccine distribution. Figure 2 shows the timing of interventions over the 18-month simulation.

**Figure 2.**
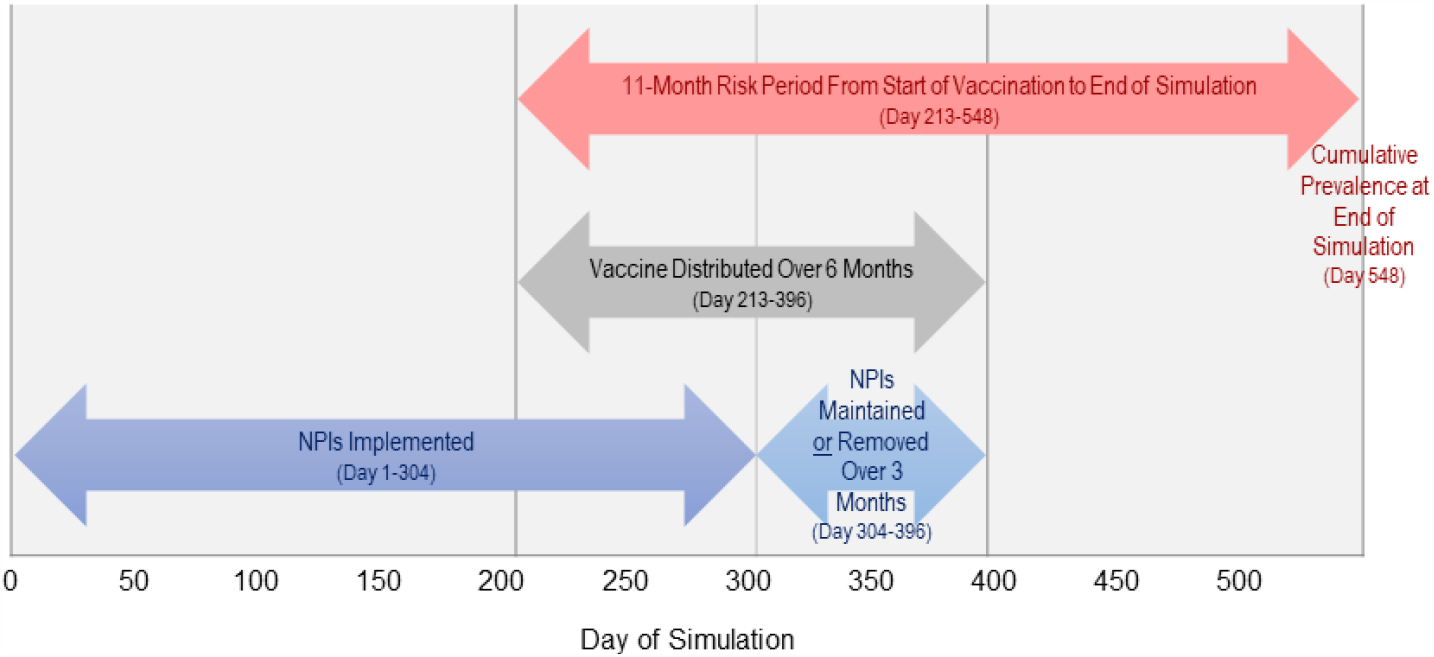
Description of Vaccination and Non-Pharmaceutical Interventions over the 18-Month Simulation.

### Outcome Analysis

The outcomes estimated included daily total infections (symptomatic and asymptomatic), hospitalizations, and deaths. For each outcome, we computed totals over the simulation period and cumulative prevalence on the final day of the simulation (Figure 2). To compare the impact of different vaccination scenarios with NPIs maintained (A1-F1) and NPIs removed (A0-F0), we computed cumulative incidences, or risks, of infection from the start of vaccine distribution and risk differences (RDs) between scenarios. The worst-case vaccination scenario with NPIs removed (F0) was designated as the referent. The mean and the standard deviation (SD) of each outcome measure were calculated across the 45 replications.

## Results

### Statewide Analysis

In a simulation of 10,490,000 people, a 50% efficacious vaccine at 25% coverage with NPIs removed (F0) resulted in almost 31% infected after 18 months (Table 1). Over the 11-month period from the onset of vaccination, 2,231,134 new infections occurred in this worst-case scenario. In contrast, the best-case 90% efficacious vaccine at 75% coverage with NPIs maintained (A1) resulted in 450,575 new infections, for a 19% absolute risk reduction. When NPIs were removed, risk reductions ranged from 9% (E0) to 18% (A0) with increasing vaccine efficacy and coverage. Scenario D0 (i.e., 50% efficacy, 75% coverage) had a greater risk reduction than Scenario C0 (i.e., 90% efficacy, 25% coverage), 13% and 8%, respectively, suggesting vaccine coverage had a greater impact on reducing infections than efficacy. Figure 3 shows more pronounced differences in daily new infections with (solid lines) and without (dotted lines) NPIs with lower vaccine efficacy and coverage.

**Table 1.**
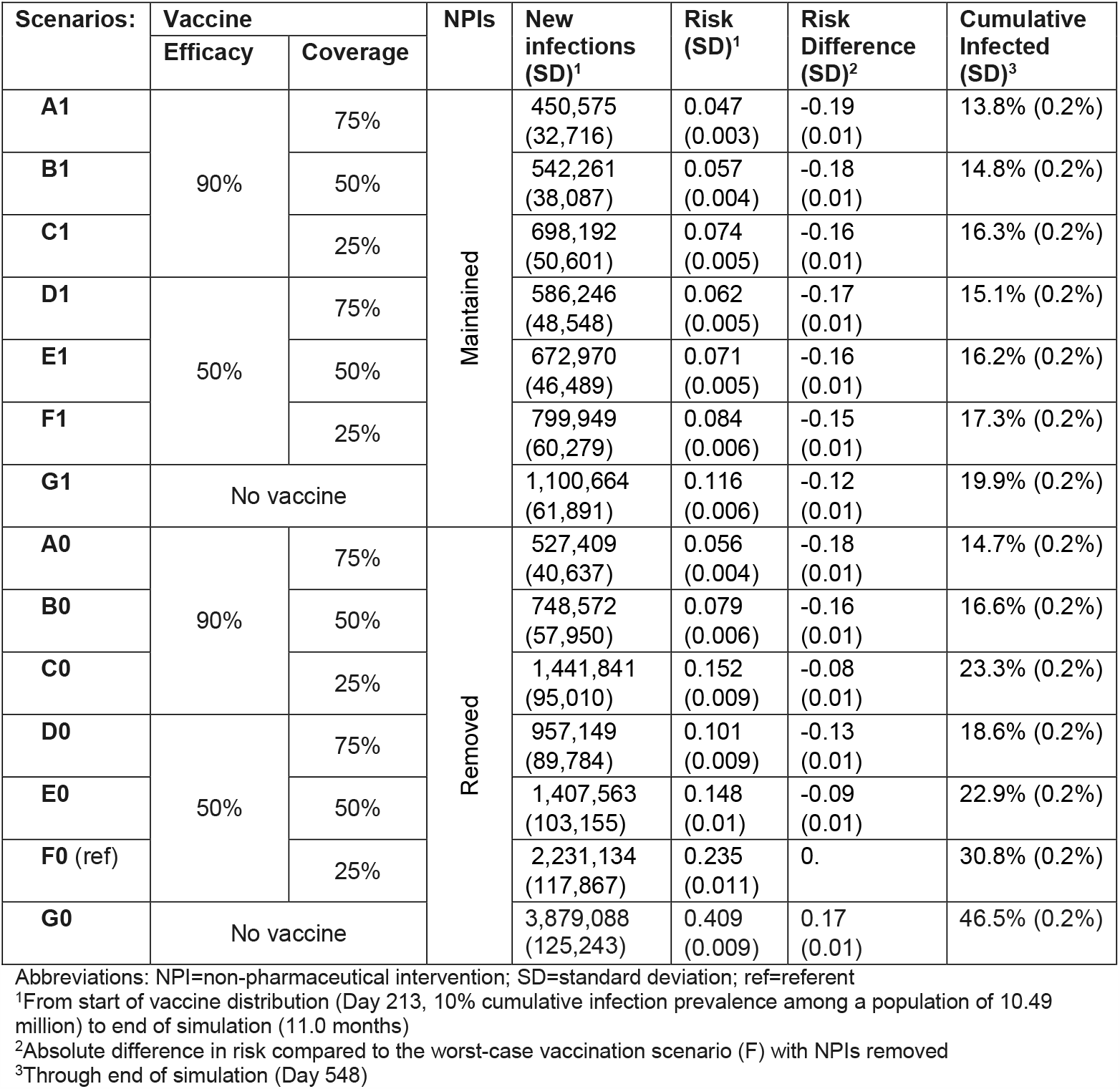
Risk of SARS-CoV-2 Infection from Start of Vaccine Distribution by Vaccination and Non-Pharmaceutical Intervention Scenarios.

**Figure 3.**
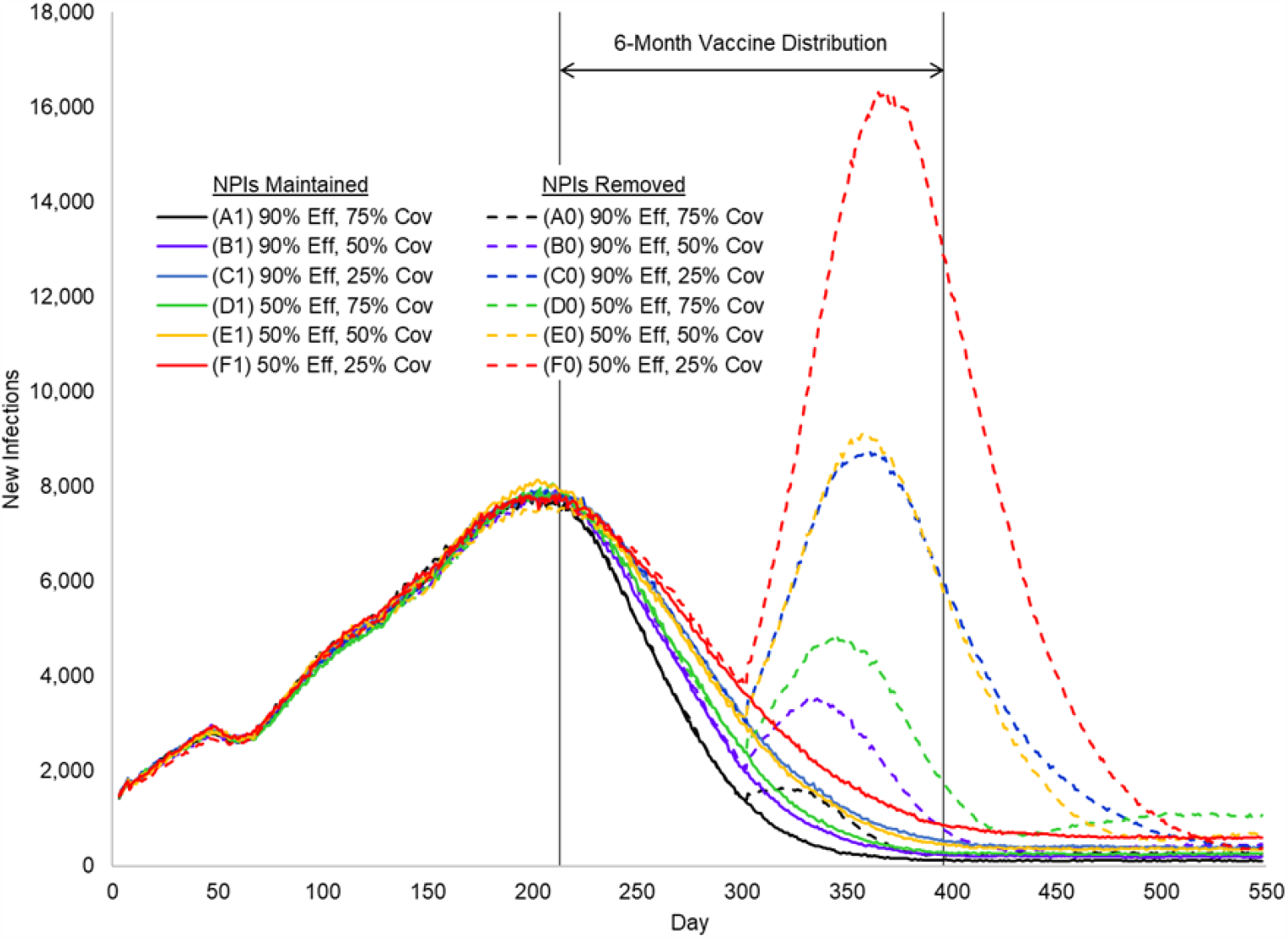
Daily New Infections by Vaccination and Non-Pharmaceutical Intervention Scenarios over the 18-Month Simulation.

All vaccination scenarios with NPIs had lower risks than without NPIs counterparts; these differences tended to increase with lower vaccine efficacy and coverage. Maintaining NPIs with the worst-case vaccination scenario (F1) reduced infections by 15% compared to this scenario without NPIs (F0). In contrast, risk reductions for Scenarios A1 and A0 were similar (19% and 18%, respectively), suggesting NPIs had a smaller impact under best-case vaccination. Similar patterns for the joint impact of vaccination and NPIs were observed for mortality risks and hospitalizations. A total 15,166 deaths (0.1% of the population) resulted from worst-case vaccination without NPIs (F0), whereas best-case vaccination with NPIs (A1) resulted in 6,789 total deaths (Table 2).

**Table 2.** COVID-19 Mortality and Hospitalizations by Vaccination and Non-Pharmaceutical Intervention Scenarios.

| Scenarios: | Vaccine |  | NPIs | New Deaths (SD)<sup>1</sup> | Mortality Risk (per 100,000) (SD) | Mortality Risk Difference (SD)<sup>2</sup> | Total Deaths (SD)<sup>3</sup> | Total Hospitalized (SD)<sup>3</sup> | Cumulative Hospitalization (per 100,000)<sup>3</sup> |
| --- | --- | --- | --- | --- | --- | --- | --- | --- | --- |
|  | Efficacy | Coverage |  |  |  |  |  |  |  |
| A1 | 90% | 75% | Maintained | 2,777 (260) | 26.5 (2.5) | -79.8 (7.0) | 6,789 (450) | 52,167 (1,086) | 497 |
| B1 |  | 50% |  | 3,227 (244) | 30.7 (2.3) | -75.6 (6.9) | 7,265 (405) | 55,924 (1,109) | 533 |
| C1 |  | 25% |  | 3,943 (301) | 37.6 (2.9) | -68.7 (7.1) | 7,981 (430) | 61,593 (1,138) | 587 |
| D1 | 50% | 75% |  | 3,364 (320) | 32.1 (3.1) | -74.2 (7.2) | 7,374 (415) | 56,852 (1,114) | 542 |
| E1 |  | 50% |  | 3,832 (310) | 36.5 (3.0) | -69.8 (7.1) | 7,935 (406) | 61,111 (1,134) | 583 |
| F1 |  | 25% |  | 4,465 (369) | 42.6 (3.5) | -63.7 (7.4) | 8,508 (442) | 65,632 (1,160) | 626 |
| G1 | No vaccine |  |  | 5,760 (405) | 54.9 (3.9) | -51.4 (7.6) | 9,721 (425) | 75,549 (1,214) | 720 |
| A0 | 90% | 75% | Removed | 3,120 (287) | 29.7 (2.7) | -76.6 (7.0) | 7,230 (463) | 55,160 (1,097) | 526 |
| B0 |  | 50% |  | 4,207 (324) | 40.1 (3.1) | -66.2 (7.2) | 8,153 (388) | 61,855 (1,128) | 590 |
| C0 |  | 25% |  | 7,510 (527) | 71.6 (5.0) | -34.7 (8.2) | 11,511 (464) | 87,656 (1,230) | 836 |
| D0 | 50% | 75% |  | 5,109 (508) | 48.7 (4.8) | -57.6 (8.1) | 9,081 (381) | 69,184 (1,154) | 660 |
| E0 |  | 50% |  | 7,282 (491) | 69.5 (4.7) | -36.8 (8.0) | 11,258 (481) | 85,290 (1,221) | 813 |
| F0 (ref) |  | 25% |  | 11,152 (683) | 106.3 (6.5) | 0. | 15,166 (589) | 116,060 (1,318) | 1,106 |
| G0 | No vaccine |  |  | 18,662 (570) | 177.9 (5.4) | 71.6 (9.2) | 22,659 (446) | 178,409 (1,473) | 1,701 |

Abbreviations: NPI=non-pharmaceutical intervention; SD=standard deviation; ref=referent
<sup>1</sup>From start of vaccine distribution (Day 213, 10% cumulative infection prevalence among a population of 10.49 million) to end of simulation (11.0 months)
<sup>2</sup>Absolute difference in risk (per 100,000) compared to the worst-case vaccination scenario (F) with NPIs removed
<sup>3</sup>Through end of simulation (Day 548)

### Subgroup Analyses

Cumulative infections, hospitalizations, and deaths by scenario varied across race/ethnicity groups (Supplemental Tables S2). Whites tended to have the fewest infections and hospitalizations, along with lower mortality. Blacks/African Americans tended to have the highest burden of infections, hospitalizations, and mortality. While Hispanics had a higher prevalence of infections compared to White, they had lower mortality (Table 3). For Scenario A1, Whites and Blacks, who had greater mortality risk in the reference scenario, also had greater reduction in mortality (−82 per 100,000) compared to Hispanics (−72 per 100,000). Differences between mortality reduction across race/ethnicity were comparable for Scenario A0 (−78/100,000 in Whites and -77/100,000 in Blacks versus - 67/100,000 in Hispanics).

**Table 3.** COVID-19 Mortality by Vaccination and Non-Pharmaceutical Intervention Scenarios Across Race/Ethnicity Groups and Urban-Rural Tracts.

| Scenarios: | Vaccine |  | NPIs | Race/Ethnicity Group |  |  |  |  |  | Rural-Urban Classification<sup>1</sup> |  |  |  |  |  |
| --- | --- | --- | --- | --- | --- | --- | --- | --- | --- | --- | --- | --- | --- | --- | --- |
|  | Efficacy | Coverage |  | White |  | Black |  | Hispanic |  | Urban |  | Suburban |  | Rural |  |
|  |  |  |  | Cum. Mort. (per 100,000) (SD)<sup>2</sup> | Mort. Diff. (SD)<sup>3</sup> | Cum. Mort. (per 100,000) (SD) | Mort. Diff. (SD) | Cum. Mort. (per 100,000) (SD) | Mort. Diff. (SD) | Cum. Mort. (per 100,000) (SD) | Mort. Diff. (SD) | Cum. Mort. (per 100,000) (SD) | Mort. Diff. (SD) | Cum. Mort. (per 100,000) (SD) | Mort. Diff. (SD) |
| A1 | 90% | 75% | Maintained | 64 (3) | -82 (6) | 73 (6) | -82 (10) | 60 (8) | -72 (14) | 62 (1) | -76 (1) | 75 (2) | -82 (4) | 72 (3) | -98 (5) |
| B1 |  | 50% |  | 68 (3) | -78 (6) | 79 (6) | -76 (10) | 64 (8) | -69 (14) | 66 (1) | -73 (1) | 78 (2) | -79 (4) | 85 (3) | -85 (5) |
| C1 |  | 25% |  | 75 (4) | -71 (6) | 86 (6) | -68 (10) | 69 (9) | -64 (15) | 72 (1) | -66 (1) | 89 (3) | -68 (4) | 88 (3) | -82 (5) |
| D1 | 50% | 75% |  | 69 (3) | -76 (6) | 80 (6) | -75 (10) | 66 (9) | -66 (15) | 67 (1) | -71 (1) | 79 (2) | -77 (4) | 85 (3) | -86 (5) |
| E1 |  | 50% |  | 75 (4) | -71 (6) | 85 (6) | -70 (10) | 70 (9) | -63 (15) | 72 (1) | -66 (1) | 88 (3) | -69 (4) | 88 (3) | -83 (5) |
| F1 |  | 25% |  | 81 (4) | -65 (6) | 90 (6) | -64 (10) | 72 (9) | -60 (15) | 77 (1) | -62 (1) | 94 (3) | -63 (4) | 98 (3) | -73 (5) |
| G1 | No vaccine |  |  | 92 (4) | -53 (6) | 102 (7) | -53 (11) | 86 (10) | -47 (16) | 88 (1) | -51 (1) | 107 (3) | -50 (4) | 112 (4) | -59 (6) |
| A0 | 90% | 75% | Removed | 68 (3) | -78 (6) | 78 (6) | -77 (10) | 65 (9) | -67 (15) | 65 (1) | -74 (1) | 82 (3) | -75 (4) | 85 (3) | -86 (5) |
| B0 |  | 50% |  | 77 (4) | -69 (6) | 87 (7) | -68 (11) | 74 (9) | -58 (15) | 74 (1) | -65 (1) | 87 (3) | -70 (4) | 91 (3) | -80 (5) |
| C0 |  | 25% |  | 110 (4) | -35 (6) | 119 (7) | -36 (11) | 99 (11) | -33 (16) | 106 (1) | -32 (1) | 123 (3) | -34 (4) | 127 (4) | -43 (6) |
| D0 | 50% | 75% |  | 86 (4) | -60 (6) | 95 (7) | -59 (11) | 81 (9) | -52 (15) | 83 (1) | -55 (1) | 83 (3) | -73 (4) | 99 (3) | -71 (5) |
| E0 |  | 50% |  | 108 (4) | -37 (6) | 115 (7) | -39 (11) | 96 (10) | -37 (16) | 102 (1) | -36 (1) | 116 (3) | -41 (4) | 121 (4) | -49 (6) |
| F0 (ref) |  | 25% |  | 146 (5) | 0 | 155 (8) | 0 | 132 (12) | 0 | 138 (1) | 0 | 157 (3) | 0 | 171 (4) | 0 |
| G0 | No vaccine |  |  | 220 (6) | 74 (8) | 226 (10) | 71 (13) | 194 (15) | 61 (19) | 206 (2) | 67 (2) | 243 (4) | 86 (5) | 260 (5) | 90 (6) |

Abbreviations: NPI=non-pharmaceutical intervention; SD=standard deviation; ref=referent
<sup>1</sup>Census tract Rural-Urban Commuting Area (RUCA) code: 1-2=urban, 3-5=suburban, and 6-10=rural
<sup>2</sup>Through end of simulation (Day 548)
<sup>3</sup>Absolute difference in cumulative mortality (per 100,000) compared to the worst-case vaccination scenario (F) with NPIs removed

Rural tracts had a higher prevalence of infections, hospitalizations, and deaths compared to urban tracts, in all scenarios (Supplemental Table S3). While urban and suburban tracts had comparable cumulative infections, suburban tracts experienced more hospitalizations and higher mortality. Mortality reduction under Scenario A1 was greatest in rural tracts (−98 per 100,000) and lowest in urban tracts (−76 per 100,000) (Table 3). For Scenario A0, the difference in mortality reduction between rural and urban tracts was smaller (−86/100,000 and -74/100,000, respectively.

## Discussion

Our study suggests that, for a population of 10.5 million, about 1.8 million infections and 8,000 deaths could be prevented with more efficacious COVID-19 vaccines, higher vaccination coverage, and maintaining NPIs, such as distancing and face mask usage. Moreover, our findings highlight the importance of continued adherence to NPIs while the population is vaccinated, particularly under scenarios of lower vaccine efficacy and coverage. Maintaining NPIs throughout the 6-month vaccine distribution period appears to reduce infections to levels seen at the beginning of the pandemic. In contrast, under scenarios with low vaccine efficacy and coverage, premature lifting of NPIs could result in a resurgence of cases with a magnitude exceeding that prior to vaccine distribution.

While initial vaccine trials have demonstrated around 95% efficacy to reduce symptomatic infections,^31^ the ability of immunization to prevent asymptomatic infection and transmission is not well understood.^3, 6, 32^ Further, protection may be limited in those who miss a booster dose or if a new strain of the virus with antibody resistance emerges.^33^Therefore, a universal vaccine efficacy of 90% modeled in our study is likely to be overly optimistic. Similarly, vaccinating at least 75% of the adult population over 6 months, as modeled in our study, will be challenging, given issues surrounding vaccine supply, distribution, and hesitancy.^34, 35^ Thus, while maintaining NPIs during vaccination efforts had less impact on preventing infections in the best-case vaccination scenario, achieving 90% efficacy and 75% coverage may not be realistic, emphasizing the need for continued NPI adherence.

Consistent with prior studies,^8, 9^ our findings suggests that higher vaccine coverage (i.e., 75% vs. 25%) could lead to a comparatively greater reduction of infections than higher vaccine efficacy (i.e., 90% vs. 50%) when NPIs are lifted. Currently, a strategy of reducing the number of doses of the mRNA vaccines to increase the vaccine supply and vaccinate more people is being debated.^36^Our study was not designed to inform decisions surrounding this vaccination policy. Although coverage is a function of vaccine supply and uptake, it was considered broadly in our study. Further, the vaccine was distributed throughout the population uniformly rather by priority groups. Comparing the scenarios with 90% efficacy and 25% coverage versus 50% efficacy and 50% coverage, in which coverage is doubled, we observed minimal differences in risk of new infections and COVID-19-associated hospitalizations and deaths.

For SARS-CoV-2, the threshold for herd immunity is estimated at 58% for an R0 of 2.4 though these values depend on population characteristics and virus transmission dynamics,^37,38^ with estimates ranging from 43% to 90%.^39, 40^ We found sufficient population immunity to slow new infections was achieved through both vaccination over 6 months and NPIs although removing NPIs during vaccination distribution resulted in additional hospitalizations and deaths. Our findings suggest coordinated efforts are needed to maximize vaccine coverage and adherence to NPIs to reduce COVID-19 burden to a level that could safely allow a resumption of many economic, educational, and social activities.

We found disparities in COVID-19 mortality by race/ethnicity that are consistent with prior findings that Blacks/African Americans are at higher risk of hospitalization and death.^41^ In our model, these disparities are partly explained by diabetes as a risk factor for hospitalization and are likely underestimated since we did not account for other high-risk conditions as well as the adverse effects of structural racism.^42, 43^ We also found higher COVID-19 hospitalizations and mortality in rural communities compared to urban communities. We did not take into account local healthcare capacity, so if hospitals or intensive care units were to become overburdened, deaths could be greater than estimated, which may be particularly relevant in rural areas. With vaccine distribution and uptake assumed to be uniform, we found notable differences in mortality reduction attributed to vaccination and NPIs across race/ethnicity groups and rurality, which can be explained by differences in risk due to age, diabetes, and mobility related to essential work. However, lower vaccine uptake among Black and rural communities due to hesitancy,^35^ and the logistical challenges of vaccines requiring ultracold storage^44^and multiple doses has the potential to further exacerbate disparities in COVID-19 mortality.

Unlike prior deterministic compartmental models of SARS-CoV-2 transmission in the context of a COVID-19 vaccine,^8-10, 45-47^ our agent-based simulation modeling allowed for heterogeneous mixing between individuals and within households, schools, workplaces, and communities and joint consideration of various intervention strategies. In this study, we did not intend to forecast COVID-19 burden for a specific population but to support public health officials by estimating the potential impact of various vaccination and NPI scenarios. For example, these results can be used by state and local health departments to inform the public on the potential effects of varying levels of vaccine as well as continued adherence to existing mitigation strategies.^48^Further, our agent-based modeling framework can be adapted to other locations using existing Census data and publicly available data on COVID-19 cases, hospitalizations, and deaths. However, this approach may be less accurate at smaller geographic regions, such as a single county, where there is a relatively small number of hospitalizations and deaths for model calibration and validation.

Our findings are conditional on our modeling assumptions and specification of vaccination and NPI scenarios. Notably, in our simulation, vaccination protects individuals from infection in an all-or-nothing manner, i.e., a proportion of susceptible individuals transition to the recovered state. However, a “leaky” vaccine considered by Bubar, et al.^45^ in which individuals can be partially protected could be more realistic. Our modeling also simplified vaccine distribution to the same period (i.e., 6 months) for all coverages analyzed. Moreover, distribution did not account for multiple doses and delayed protection, was uniform across the population, and was not prioritized by subpopulations^2, 44^ (e.g., older versus younger people). Lastly, although vaccine coverage is a function of supply and uptake, we considered it broadly in this study. Our ongoing work models alternative protective effects of multiple COVID-19 vaccines, the phased allocation of vaccines to priority populations (e.g., essential workers, elderly), and potential limited coverage due to supply chain issues and vaccine hesitancy. We will incorporate emerging evidence on vaccine efficacy to reduce symptomatic illness and transmission and epidemiologic studies of seroprevalence and reinfection into our model structure and parameters.

In addition to the modeling assumptions discussed above, there are notable limitations to our study. The unique population characteristics for the studied state, including population demographics, prevalence of diabetes, rurality, mobility and adherence to NPIs, influence the transmission of infections and COVID-19 morbidity and mortality. General insights into the comparative and joint impact of vaccination and NPIs may apply to other states or regions though exact numerical values will vary. The broad age category for adults (20-64 years old) limits age-specificity of outcomes within this range. Demographic attributes (e.g., age and race/ethnicity) within census tract were assigned independently, and interactions were not fully captured. Our model framework assumed only hospitalized individuals can transition to death, so mortality estimates of the early phase of the pandemic may be inaccurate since many deaths occurred in nursing homes and outside the hospital. Lastly, while the variability in outcomes across replications was computed, our results do not reflect uncertainty in the model structure and parameter specification.

Our study suggests vaccinating the majority of the adult population and continued adherence to NPIs such as physical distancing and face mask usage will have the greatest impact on ending the current COVID-19 pandemic. These findings emphasize the need for resources and coordination to achieve high vaccine coverage and continued adherence to NPIs before safely resuming many pre-pandemic activities. Future research will incorporate the continually growing evidence on the efficacy of COVID-19 vaccines and the epidemiology of the pandemic.

## Data Availability

The manuscript utilizes publicly available data sources and model parameters from the literature. All sources are cited in the manuscript.

## Acknowledgements

The authors thank Paul Delamater and Mark Holmes for their contributions to the COVID-19 Simulation Integrated Modeling (COVSIM) initiative.

## Supplement 1: Data Sources and Disease Model

In this document, we provide additional information on the model used to project the spread of COVID-19 infections and associated hospitalizations and deaths. The model was based on the structures of ones previously published for influenza [1-3] and COVID-19 [4-6] pandemics.

### Agent-Based Network

A network of agents allowed for interactions and transmission in households, workplace and school peer groups, and communities of census tracts. The model used 1,017,720 agents to represent a statewide population of 10,490,000. Data values to generate individual agents were drawn from the US Census and were grouped into households. Census tract data was used to determine the size of the household (1 to 6), whether or not children were present [7, 8], and the race/ethnicity of the household (with reporting for Black Only, White Only, Hispanic not White or Black, etc.) For the simulation, we assumed all individuals in the household are the same race/ethnicity and that it is independent of other factors. This last assumption was based on the census tract tables that were publicly available for recent years; it implies that our model is capturing less of the heterogeneity than exists in reality. Individual household members were assigned to age in five categories (age 0 to 4; age 5 to 9; age 10 to 19; age 20 to 64; age 65 and greater) [7], with the head of the household always 18 or older. As an indicator for high risk conditions, we used statewide prevalence of diabetes by race/ethnicity and adjusted for age [9]. The conditional probability of an agent having diabetes given their age and race can be calculated from the statewide prevalence values. While there are other conditions, diabetes affects many people and has a large impact on hospitalizations. During the day, all agents 5 to 19 interacted in peer groups (“schools”), and agents 19 to 64 interacted in peer groups (“work”) according to commuting patterns. The latter was determined from workflow data indicating the people who live in a census tract and the percentage who work in each of the other census tracts [10]. When schools are closed (or hybrid), children stay at home (or stay home every other day). All agents interacted with household members at night.

For adult agents age 20 to 64, working from home or outside of the house was determined by time-varying mobility factors. Specifically, we used mobility data generated by SafeGraph, a data company that aggregates anonymized location data from numerous applications in order to provide insights about physical places, via the Placekey Community (Placekey.io) [11]. To enhance privacy, SafeGraph excludes census block group information if fewer than five devices visited an establishment in a month from a given census block group. SafeGraph tracks device location data and classifies each as working full time if that device spent greater than 6 hours at a location other than their home geohash-7 during the period of 8 am - 6 pm in local time. The data indicate the count of devices classified as working full-time. As in the publicly available Google mobility data [12], we computed the ratio of devices at home in comparison to the month of January to quantify the proportion of working-age adults staying home. Because of sparseness of data, we grouped points at the census tract level stratified by urban, rural, or suburban according to the Rural-Urban Commuting Area (RUCA) code and whether the tract was in the 1st, 2nd, 3rd, or 4th quartile of household median income for the state level. Beginning in month 6 we assume that the mobility factors stay the same. Based on the mobility data from SafeGraph and Google on mobility in contexts outside of work, we assume that the community mobility follows the same pattern as the workplace but to a lesser extent. For example, if the mobility is at 60% then the community factor is 88% of the original community value (specific calculation for new Community parameter: (1-((1-0.6)/3.5) * (community_parameter = 0.23) = 0.204). A reduction in the number of people in the community is modeled by a reduction in the community infectious hazard. In the workplace, the reduction of people follows the workplace mobility values. This decrease in workplace activity, reduces the workplace infectious hazard. Because of sparseness of data in rural areas, we grouped SafeGraph data at the census tract level stratified by urban, rural, or suburban according to the RUCA classification system and whether the tract was in the 1st, 2nd, 3rd, or 4th quartile of household median income for the state level (Figure S1). To validate this approach, we compared to the Google mobility data available at the county level [12].

**Figure S1.**
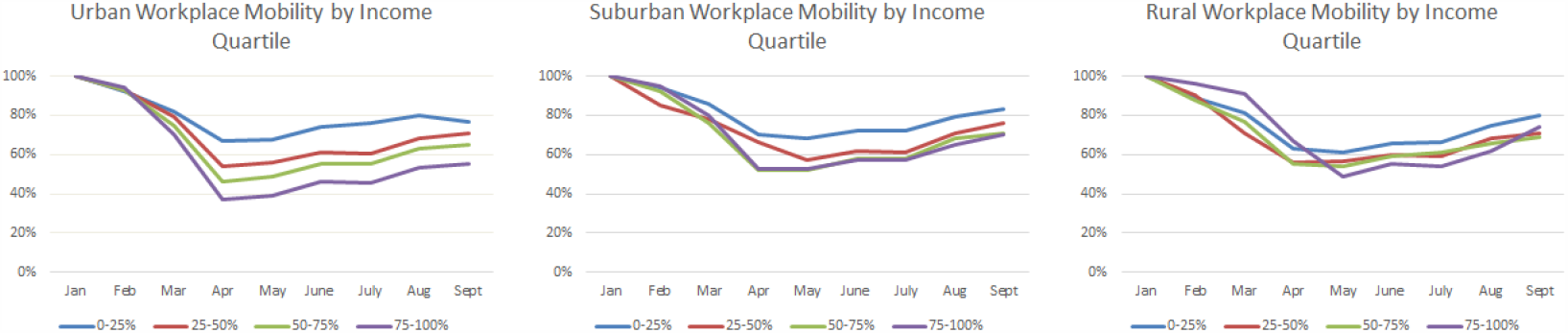
Time-Varying Workplace Mobility Estimates by Census Tract Urban-Rural Classification and Median Household Income Quartile.

### Disease Progression within an Individual

The underlying model of disease progression was assumed to be a variant of a Susceptible-Exposed-Infectious-Recovered (SEIR) model. At a given point in time, each individual is in exactly one of the following states: susceptible (S), exposed (E), pre-symptomatic (IP), asymptomatic (IA), symptomatic (IS), hospitalized (H), recovered (R), or dead (D). The list of input parameters and associated references is provided in Table S1. Additional details are available in the supplement of Keskinocak, et al. [5].

Several parameters determine the length of time within each state in the SEIR including the mean and standard deviation on the time before an exposed patient becomes pre-symptomatic, the average length of time of the pre-symptomatic phase, the distribution of time within the symptomatic (S) stage, the distribution of the length of hospitalization, and the ratio of the duration of the symptomatic and asymptomatic states. Parameters related to the transmission between states include the probability of moving to symptomatic (from IP), the probability of hospitalization (from IS), and the probability of death (from H); probabilities out of a state must sum to 100%. The probability of hospitalization or death is age-specific. The probability of hospitalization is also specific to whether someone has diabetes or not, matching both the relative risk and the overall hospitalization rate [13, 14]. The overall Infection Fatality Rate (i.e., the probability that an exposed person will die) as quantified from the simulation of transitions is approximately 0.46%, which is consistent with estimates of IFR estimated by Hauser, et al. [15], which was also used for the CDC Pandemic Planning Scenarios.

The infectivity of the virus at the beginning of the outbreak without interventions is summarized by basic reproductive number R0, and the transmission rate (denoted as β). The proportion of transmissions that occur at either the IP or IA stage is θ, and the proportion of infections generated by individuals who are never symptomatic is ω. In the absence of interventions, the proportion of transmission that occurs outside households is γ, and the proportion of transmission outside households that occur in the community is δ.

One of the important differences in comparison to the values used for influenza [1], is that the proportion of transmissions that can occur by people without symptoms is much higher. In comparison to the values used for the state of Georgia (4), δ is a little higher. For this paper we also take the import rate of cases to be lower (45 compared to 100) based on factors such as the airport size, commuting in or out of the state, etc.

**Table S1.** Key Model Parameters.

| Parameter | Estimates | References |
| --- | --- | --- |
| Exposed (E) Duration | Weibull with mean 4.6 days | [16, 17] |
| Pre-symptomatic (IP) Duration | 0.5 days | [16] |
| Hospitalized (H) Duration | Exponential with mean 10.4 days | [16, 18] |
| Symptomatic (S) Duration | Exponential with mean 2.9 days | [19] |
| Symptomatic-Asymptomatic Duration Ratio | 1.5 | [16] |
| Probability of Symptomatic (from IP) | 0.50-0.82 | [20-24] |
| Probability of Hospitalization without Diabetes | 0.005 for age 0-19;<br>0.06 for age 20-64;<br>0.1 for age 65+ | [13, 14] |
| Probability of Hospitalization with Diabetes | 3 times rates above |  |
| Probability of Death (from H) | 0 for age 0-19;<br>0.0515 for age 20-64;<br>0.3512 for age 65+ | [13, 14] |
| $R_0$ | 2.4 | [25-27] |
| $\beta$ transmission rate | 1.12 | [26] |
| $\theta$ (probability IP to IA) | 0.48 | [28] |
| $\omega$ (proportion infections by IA) | 0.24 | [28] |
| $\gamma$ (proportion of transmission that occur outside households) | 30% | [1, 5] |
| $\delta$ (proportion of infections outside households that occur in community) | 0.23 | [5]; Calibration |
| FlatImportRate | 45 | [5]; Calibration |
| Infection Fatality Rate | 0.46% | Results from model transitions with other parameters |

### Stochastic Simulation across Network

To seed infections in agents, the confirmed cases at the county level over time were collected from data hosted by The New York Times [29]. Initial infections (confirmed cumulative cases by March 24 times 10 [30]) were assigned to census tracts randomly according to the population within each tract in the county using the Huntington-Hill method.

The primary sources of randomness for the simulations include four types: (i) the structure of network, i.e., the random assignment of agents to households and peer groups (ii) the individual agents who are initially infected with the seed infections; (iii) whether an infected individual will transmit the virus to another person in the household, peer group, and/or community at a point in time; and (iv) the duration within a disease state. For each scenario, 45 replications of the simulation were run. The mean and standard deviation of each outcome was computed across the simulation replications. The number of replications was chosen so the mean values between scenarios have low variability until vaccine distribution begins.

### Validation

The simulation was calibrated and validated on cumulative infections, hospitalizations, and deaths [31, 32]. Deaths and hospitalizations are appropriate for direct validation; while not perfect, they are a reasonable estimate of true hospitalizations and deaths. However, it is well-recognized that the true cases are much greater than those that are lab-confirmed, and this value has changed over time (10-12 times [30] then later 8 times [33]). Using the Infection Fatality Rate (IFR) of 0.5% [15] and the average time from exposure to death (21 days), we computed the daily expected number of true cases that would have been needed to result in the recorded deaths. The average daily ratio of computed cases to lab-confirmed cases (or the “lab-multiplier”) ranges from approximately 10 initially to 4 times (after 147 day). The estimates obtained in this way for one state and nationally are also consistent with CDC published accounts of antibody testing to identify the number of true cases [33]. This approach does not take into account the changing age-distribution of positive cases, which likely affected the timing of deaths. We compare our simulated results to actual deaths and hospitalizations and cases adjusted by the lab multiplier. For the baseline scenario without vaccine, our estimates are within 6% of the reported cumulative values.

### Simulated Interventions

The following interventions were analyzed as scenarios defined in the main text:

- <u>Vaccination:</u> Vaccine efficacy and the percentage of adults (20 year and older) receiving the vaccine (“coverage”) determined the impact of vaccination.
  - Efficacy is defined as the percentage of those vaccinated who transition to recovered if they were exposed. This implies that the vaccine can prevent both the symptomatic and asymptomatic states. For future research, we also developed a vaccine effect that shifts symptomatic infections to asymptomatic.
  - Coverage is defined as the percent of the adult population who received the vaccine. Starting when the cumulative infection prevalence for the entire population was 10%, randomly selected adult agents (20 years or older) could be vaccinated regardless of disease state. The coverage parameter was reached over a 6-month distribution period. Based on expected vaccine supply, we assumed 18% of possible vaccinations occurred in months 0-2, 32% of possible vaccinations occurred in months 2-4, and 50% of vaccinations occurre in months 4-6. During each 2-month interval, the same amount of vaccine was distributed each week.
- <u>Non-Pharmaceutical Interventions (NPIs)</u>
  - Pre-Vaccine
    ▪ Mobility was modeled using SafeGraph mobility data. Applying these data modeled the reduction in peer group activity for adults in the workplace and for all agents in the community setting. In NC, various orders were specified when the population should stay at home as much as possible including non-essential workers. We are capturing this through the realized mobility data over the time period studied. (See discussion above on agent mobility.) We assume that there are no additional orders that change the mobility after September.
    ▪ Voluntary quarantine - if an agent had a family member that was symptomatic then the agent voluntarily quarantined until the agent has recovered. The probability of this behavior was higher during the first 50 days of the simulation. For the first 46 days the probability of voluntary quarantine was 0.3. At day 46 it was raised to 0.6. After day 46 it decreased by 0.15 per week until reaching 0.15. It remained at 0.15 for the duration of the simulation.
    ▪ Schooling - Until August 24, 2020, no agents were attending school. After August 24, school-age agents attended school following a hybrid schooling policy [34]. In this policy, the agents were split into two groups. If the day of the simulation was odd, then half of the agents attended school, and if the day was even, then the other half attended school. Attending school corresponds to setting the agents peer group to active.
    ▪ Masks were applied uniformly throughout the population. Masks were worn in the community and peer group settings, not the household. The effectiveness of masks was controlled by two parameters: adherence and efficacy. Adherence refers to the proportions of the agents that wear masks. Efficacy refers to the ability of masks to reduce susceptibility and infectivity. If an agent wore a mask they become less likely to become exposed to the disease, and less likely to spread the disease. Once an agent is assigned a mask, the assignment did not change.
  - Intervention removal occurs exactly halfway through the 6-month vaccine distribution (i.e. 3 months after distribution begins)
    ▪ Probability of voluntary quarantine was set to 0 immediately.
    ▪ The schooling returned to normal. All of the children agents’ peer groups were set to active and agents that are not symptomatic attend school. There was not a gradual increase; full school activity resumed immediately.
    ▪ Workplace peer groups returned to normal. All of the agents that could work began attending work. All of the adult agents’ peer groups were set to active. There was not a gradual increase; work place activity resumed immediately.
    ▪ Community interaction at the census tract level returned to normal level immediately.
    ▪ Random individuals removed masks as the vaccine was distributed equaling the number of people that had been vaccinated. At the end of the vaccine distribution period, the same number of individuals vaccinated were no longer using face masks.

## Supplement 2: Supplemental Results

**Table S1.**
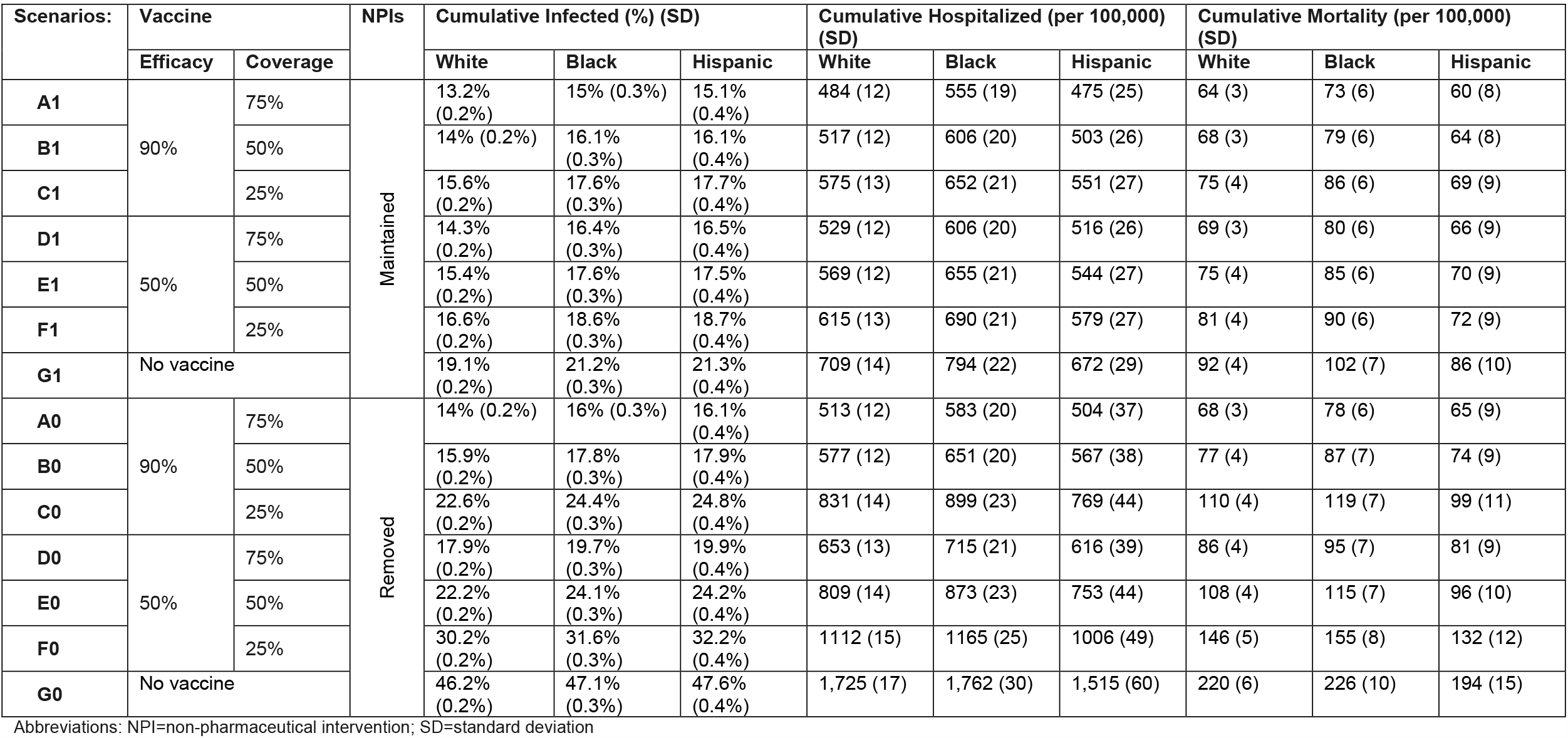
Cumulative infections, hospitalizations, and deaths by vaccination-NPI scenario across race/ethnicity groups.

**Table S2.** Cumulative infections, hospitalizations, and deaths by vaccination-NPI scenario across rural-urban classification^1^.

| Scenarios: | Vaccine |  | NPIs | Cumulative Infected (%) (SD) |  |  | Cumulative Hospitalized (per 100,000) (SD) |  |  | Cumulative Mortality (per 100,000) (SD) |  |  |
| --- | --- | --- | --- | --- | --- | --- | --- | --- | --- | --- | --- | --- |
|  | Efficacy | Coverage |  | Urban | Suburban | Rural | Urban | Suburban | Rural | Urban | Suburban | Rural |
| A1 | 90% | 75% | Maintained | 13.7% (0.1%) | 13.9% (0.2%) | 14.6% (0.2%) | 488 (4) | 519 (9) | 541 (12) | 62 (1) | 75 (2) | 72 (3) |
| B1 |  | 50% |  | 14.6% (0.1%) | 14.7% (0.2%) | 16.1% (0.2%) | 522 (4) | 549 (9) | 604 (12) | 66 (1) | 78 (2) | 85 (3) |
| C1 |  | 25% |  | 16.2% (0.1%) | 16.3% (0.2%) | 17.5% (0.2%) | 576 (4) | 608 (9) | 650 (12) | 72 (1) | 89 (3) | 88 (3) |
| D1 | 50% | 75% |  | 15% (0.1%) | 14.8% (0.2%) | 16.2% (0.2%) | 531 (4) | 554 (9) | 611 (12) | 67 (1) | 79 (2) | 85 (3) |
| E1 |  | 50% |  | 16% (0.1%) | 16.3% (0.2%) | 17.3% (0.2%) | 569 (4) | 614 (9) | 647 (12) | 72 (1) | 88 (3) | 88 (3) |
| F1 |  | 25% |  | 17.1% (0.1%) | 17.4% (0.2%) | 18.6% (0.2%) | 613 (4) | 649 (9) | 696 (12) | 77 (1) | 94 (3) | 98 (3) |
| G1 | No vaccine |  |  | 19.7% (0.1%) | 19.8% (0.2%) | 21.5% (0.2%) | 704 (4) | 747 (10) | 811 (13) | 88 (1) | 107 (3) | 112 (4) |
| A0 | 90% | 75% | Removed | 14.6% (0.1%) | 14.6% (0.2%) | 16% (0.2%) | 514 (4) | 547 (9) | 589 (12) | 65 (1) | 82 (3) | 85 (3) |
| B0 |  | 50% |  | 16.5% (0.1%) | 16.3% (0.2%) | 17.6% (0.2%) | 580 (4) | 605 (9) | 646 (12) | 74 (1) | 87 (3) | 91 (3) |
| C0 |  | 25% |  | 23.4% (0.1%) | 22.1% (0.2%) | 24.5% (0.2%) | 828 (4) | 823 (10) | 915 (13) | 106 (1) | 123 (3) | 127 (4) |
| D0 | 50% | 75% |  | 18.5% (0.1%) | 18.1% (0.2%) | 19.7% (0.2%) | 677 (4) | 706 (10) | 775 (12) | 83 (1) | 83 (3) | 99 (3) |
| E0 |  | 50% |  | 22.9% (0.1%) | 22.1% (0.2%) | 24% (0.2%) | 803 (4) | 822 (10) | 886 (13) | 102 (1) | 116 (3) | 121 (4) |
| F0 |  | 25% |  | 30.9% (0.1%) | 29.2% (0.2%) | 32.3% (0.2%) | 1099 (4) | 1083 (11) | 1203 (14) | 138 (1) | 157 (3) | 171 (4) |
| G0 | No vaccine |  |  | 46.3% (0.1%) | 45.4% (0.1%) | 50.1% (0.2%) | 1674 (5) | 1711 (12) | 1901 (15) | 206 (2) | 243 (4) | 260 (5) |

Abbreviations: NPI=non-pharmaceutical intervention; SD=standard deviation
<sup>1</sup>Census tract rural-urban commuting area (RUCA) code: 1-2=urban, 3-5=suburban, and 6-10= rural

